# TOWARDS A SINGLE PARAMETER FOR THE ASSESSMENT OF EEG OSCILLATIONS

**DOI:** 10.1101/2022.02.06.22270548

**Authors:** Arturo Tozzi, James F. Peters

## Abstract

The single macroscopic flow on the boundary of a closed curve equals the sum of the countless microscopic flows in the enclosed area. According to the dictates of the Green’s theorem, the counterclockwise movements on the border of a two-dimensional shape must equal all the counterclockwise movements taking place inside the shape. This mathematical approach might be useful to analyse neuroscientific data sets for its potential capability to describe the whole cortical activity in terms of electric flows occurring in peripheral brain areas. Once mapped raw EEG data to coloured ovals in which different colours stand for different amplitudes, the theorem suggests that the sum of the electric amplitudes measured inside every oval equals the amplitudes measured just on the oval’s edge. This means that the collection of the vector fields detected from the scalp can be described by a novel, single parameter summarizing the counterclockwise electric flow detected in the outer electrodes. To evaluate the predictive power of this parameter, in a pilot study we investigated EEG traces from ten young females performing Raven’s intelligence tests of various complexity, from easy tasks (n=5) to increasingly complex tasks (n=5). Despite the seemingly unpredictable behavior of EEG electric amplitudes, the novel parameter proved to be a valuable tool to to discriminate between the two groups and detect hidden, statistically significant differences. We conclude that the application of this promising parameter could be expanded to assess also data sets extracted from neurotechniques other than EEG.

## INTRODUCTION

The circulation of flows around a two-dimensional closed curve can be described by the integral of a two-dimensional vector field which exemplifies the “macroscopic” circulation around the boundary of the closed curve. In turn, the “microscopic” circulation inside the enclosed surface can be depicted in terms of numerous small, closed curves which represent the vector field’s propensity to circulate at that location. It is noteworthy that the microscopic flows match the curl i.e., a vector operator able to define the infinitesimal circulation of vector fields in three-dimensional spaces (Schey 1997; Vines et al., 2021). Here the Green’s theorem from vector calculus comes into play, suggesting that the sum of all the microscopic circulations occurring in the area delimited by the closed curve equals the macroscopic circulation occurring on the curve’s edge (Green, 1828; Cauchy 1846; Arfken 1985). The Green’s theorem holds only for curves oriented counterclockwise, such that the border is defined as a positively oriented boundary surrounding the two-dimensional region.

In this paper we provided an effort to assess neuroscientific issues in terms of the powerful theoretical apparatus dictated by the Green’s theorem. In particular, we focused on two-dimensional data sets extracted from EEG traces during Raven’s intelligence tests of various complexity, from easy tasks to difficult tasks. Once achieved the electric oscillations extracted from the EEG traces, we used the Green’s theorem which gives us the opportunity to use a single parameter to describe the nervous dynamics. We suggest that this novel parameter could be useful to describe at least part of the brain electric activity when the latter is assessed not just through EEG, but also through other neurotechniques such as e.g., fMRI and magnetoencephalography.

## PATIENTS AND METHODS

### EEG data

We analysed EEG traces of young female students (n=10, mean age: 21 years) that were presented a non-verbal test of Raven’s Advanced Progressive Matrices Plus used to quantify general human intelligence and abstract reasoning (Mazhirina et al., 2016; Zhuo et al., 2021). Half of the subjects (n=5) were assessed with less difficult items (henceforward Raven Easy), while the other half (n=5) were assessed with increasingly complex items (henceforward Raven Difficult). The procedures are described by Jaušovec and Jaušovec (2005 and 2010), accomplished according to the Declaration of Helsinki and approved by the Ethics Committee of the University of Maribor, Slovenia. The EEG was recorded while students were performing the two different tasks, using a Quick-Cap with sintered (Silver/Silver Chloride; 8mm diameter) electrodes. The EEG activity was recorded over nineteen scalp locations (i.e., Fp1, Fp2, F3, F4, F7, F8, T3, T4, T5, T6, C3, C4, P3, P4, O1, O2, Fz, Cz and Pz) according to the International Federation’s Ten-twenty Electrode Placement System. A ground electrode was applied to the forehead and ll leads were referenced to linked mastoids (A1 and A2). The bandpass of the digital EEG data acquisition and analysis system (SynAmps) was 0.15-100.0 Hz. The voltage gain was approximately –6dB at cutoff frequencies. Every EEG trace was digitized online at 1000 Hz with a gain of 1000 (resolution of 084 μV/bit in a 16 bit A to D conversion) and stored on a hard disk. All epochs showing amplitudes above +/-50 microV (<3%) were automatically removed from computations to avoid the occurrence of artifacts of the visualizing algorithm while plotting the EEG power maps. The raw data extracted from the experimental EEG activity, i.e., the matrices displaying the electric values in μV/milliseconds for every cortical area, were plotted in terms of a series of two-dimensional coloured ovals, different colours standing for different amplitudes (**Figure 1A**). Every oval illustrates a one-millisecond time frame.

**Figure 1.**
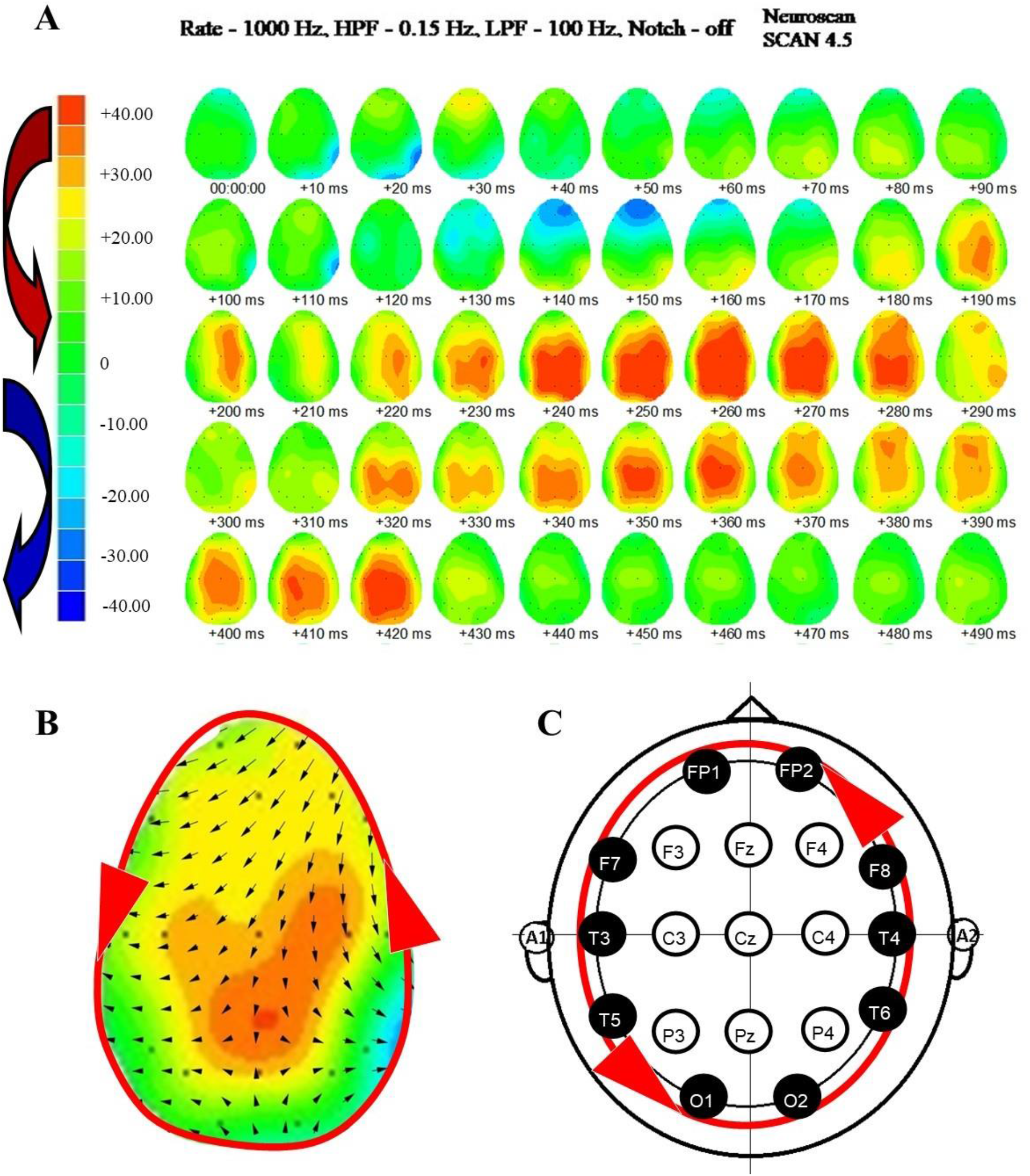
EEG traces and the corresponding electric flows. **Figure 1A**. Fifty ovals illustrate a 500 milliseconds, two-dimensional reconstruction of EEG waves from a subject performing a Raven Easy task. The different colours measure different detectable electric amplitudes (in μV) across different brain areas. The positive flows are conventionally deemed to occur anticlockwise (red curved arrow), while the negative flows clockwise (blue curved arrow). Although the EEG has been recorded using time windows of one millisecond, the Figure shows just the images taken every 10 milliseconds. **Figure 1B**. Magnification of a single frame displaying a hypothetical vector field’s grid (black arrows inside the oval). According to the Green’s theorem, the counterclockwise flow (red circle with arrows) surrounding the EEG trace’s boundary equals all the flows taking place on the enclosed vector field. **Figure 1C**. Once the counterclockwise circle is projected to a standard EEG mounting, the Green’s theorem dictates that the flow taking place on the boundary can be measured by looking just at the external electrodes (highlighted in black).

### The Green’s theorem for the appraisal of EEG data

As stated above, the electric flows extracted from EEG data can be illustrated as front waves of different colours taking place on two-dimensional ovals, the latter summarizing the scalp locations of the EEG electrodes. In touch with the Green’s theorem’s counterclockwise requirement, we conventionally regarded the EEG positive amplitudes as taking place anticlockwise and the EEG negative amplitudes as taking place clockwise (**Figure 1A**). The next step was to describe the electric amplitudes in every EEG oval frame in terms of a vector field, i.e., a quantity displaying magnitude and direction (angle) at every particular location (**Figure 1B**). Using the Green’s theorem’s dictates to assess EEG data sets, we may state what follows: the sum of the amplitudes detectable inside an oval (i.e., the vector field described in **Figure 1B**) must equal the values of amplitude detected on the surface of the same oval (i.e., the counterclockwise flow taking place on the oval’s surface is illustrated by the red path in **Figure 1B)**. This means that a single flow detectable on the border of an oval timeframe is capable of operationally summarize the whole electric oscillations detectable in the enclosed surface.

Once established that the flows on the surface of two-dimensional manifolds can be assessed in terms of the flows taking place of their edge, our next step was to operationally find the appropriate neuroscientific counterpart. The flows taking place on the oval’s edge correspond to the electrodes lying on the peripheral, outer borders of the EEG mounting (**Figure 1C**). Therefore, we evaluated the values of all the electric amplitudes detectable in the following peripheral scalp locations: Fp1, Fp2, O1, O2, F7, F8, T3, T4, T5 and T6, while the central electrodes F3, F4, T3, T4, T5, T6, C3, C4, P3, P4, Fz, Cz and Pz were excluded from our analysis (**Figure 1D**).

This means that for every timeframe (corresponding to 1 millisecond) we summed the values of the electric amplitudes experimentally detected in the outer locations Fp1, Fp2, O1, O2, F7, F8, T3, T4, T5, T6 and achieved μV/milliseconds plots. The sum can be expressed by a single value, i.e., a novel parameter able to summarize the EEG electric activity of the brain. We compared the values of this single parameter achieved from the subjects undergoing the Raven Easy and Raven Difficult tests. An independent two-sample, unpaired Student’s t test was used to determine if the means of two sets of data were significantly different from each other.

## RESULTS

The patients were clustered in two groups: Easy and Difficult Raven test. The amplitudes measured in the outer EEG electrodes (henceforward outer amplitudes) were summed and plotted in μV/milliseconds for a time length of 500 milliseconds. To provide an example, **Figure 2A** illustrates the plot extracted from one of the subjects attending a Raven Easy test. Each point of the line represents the sum of the amplitudes detected every millisecond in the peripheral scalp locations Fp1, Fp2, O1, O2, F7, F8, T3, T4, T5, T6.

**Figure 2A.**
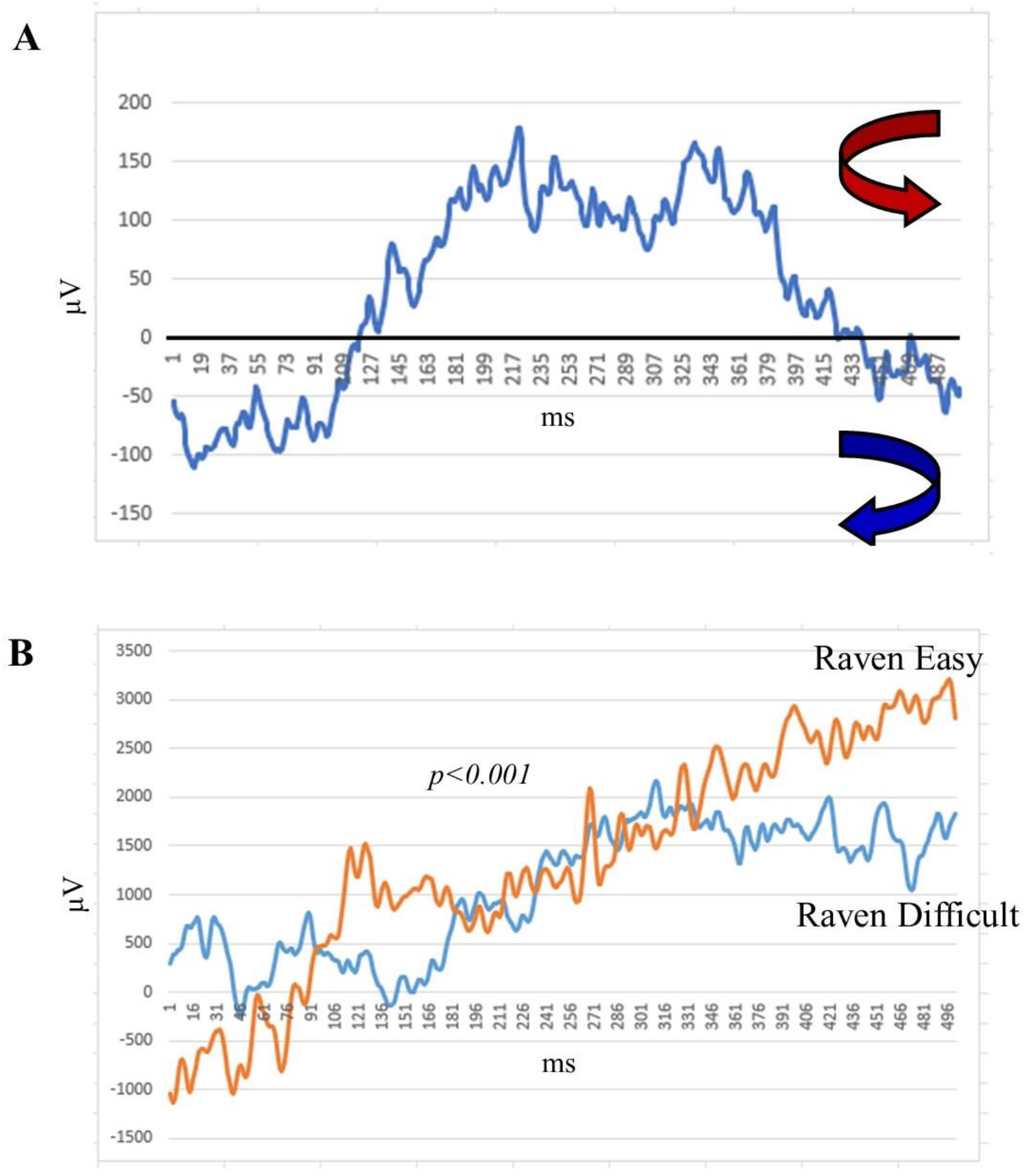
The plot (μV/milliseconds) depicts the sum of the outer amplitudes, i.e., the amplitudes detected in the locations corresponding to the electrodes Fp1, Fp2, O1, O2, F7, F8, T3, T4, T5, T6 during a Raven Easy task. The figure is drawn from the data of the coloured EEG plot illustrated in Figure 1A. **Figure 2B**. Comparison of two plots summarizing the outer amplitudes mean in the five Raven Easy and in the five Raven Difficult tests.

The mean ± SD of outer amplitude was 267,7414244 ± 521,106 μV for the patients who underwent the Easy Raven test, while 213,7800816 ± 266,8129 μV for the patients who underwent the Difficult Raven test. The Student’s t test showed that the means of two sets of outer amplitudes (Raven Easy vs. Raven Difficult) were significantly different from each other (p<0.001). Therefore, significant differences in outer amplitude oscillations related to different test difficulty were observed. It is noteworthy that the soaring values of SD suggest a huge variability in the EEG amplitude values. Despite such seemingly unpredictable behavior of the electric amplitudes, the novel, single parameter which summarizes the outer amplitudes of the peripheral electrodes proved to be a very valuable tool. Indeed, the novel parameter enabled hidden statistically significant differences to be detected, leading to a powerful operational distinction between the two groups under examination.

## DISCUSSION

The Green’s theorem says that every counterclockwise path on the border of a closed line conveys the vector fields’ behaviour taking place inside the area surrounded by the border. The two-dimensional Green’s theorem and its three-dimensional counterpart, i.e., the Stoke’s theorem, have been widely used in different scientific fields. To provide an example, they were used to assess the relationships between the charges and the scalar potentials’ horizon-values in black holes (Gibbons and Perry, 1978; Heusler 1998), suggesting the intriguing possibility to evaluate the internal flows of black holes by means of the holographic principle applied to their surfaces. In this paper, we used the Green’s theorem to describe the electric activity of the human brain in terms of the flows taking place on particular locations of the EEG mounting, i.e., the peripheral, outer electrodes. We found that a single parameter is able to summarize the general activity of the brain and to find statistically different outcomes in populations performing different cognitive operations. Even though we chose to focus on the specific task of the human intelligence, every experimentally detectable cognitive task can be assessed through the Green’s theorem, from visual perception to memory, from intention to decision making, and so on (Cisek 2019). We evaluated the relatively short time windows of 500 milliseconds in an effort to detect the very initial changes in brain activity during intelligence tasks and predict next-to-come, longer latency behaviours. Nevertheless, every technically achievable time window (either longer or even shorter) can be explored through the Green’s theorem. We chose to focus on EEG traces since we had at our disposal the rough data sets available from the published papers of the lamented Norbert Jausovec. However, the Green’s theorem can be used to investigate the nervous flows and oscillatory wave fronts achieved from different neurotechniques, such as, e.g., fMRI, magnetoencephalography and so on.

Our description of counterclockwise flows taking place in two-dimensional nervous traces can be extended also to the description of their three-dimensional counterparts. While the Green’s theorem depicts the two-dimensional case, the above mentioned Stoke’s theorem illustrates the three-dimensional case (Pontryagin 1959). In **Figure 3A**, a three-dimensional EEG headmodel is illustrated. For the right-hand rule of vector calculus, the length of every vector (the red arrows in Figure) summarizes the microscopic curl activity taking place in the corresponding scalp location. Note that some arrow turns clockwise, while others counterclockwise (not illustrated in Figure). For the Stoke’s theorem, the counterclockwise activity taking place on the EEG border equals the sum of all the counterclockwise activities depicted by the red vectors.

**Figure 3A.**
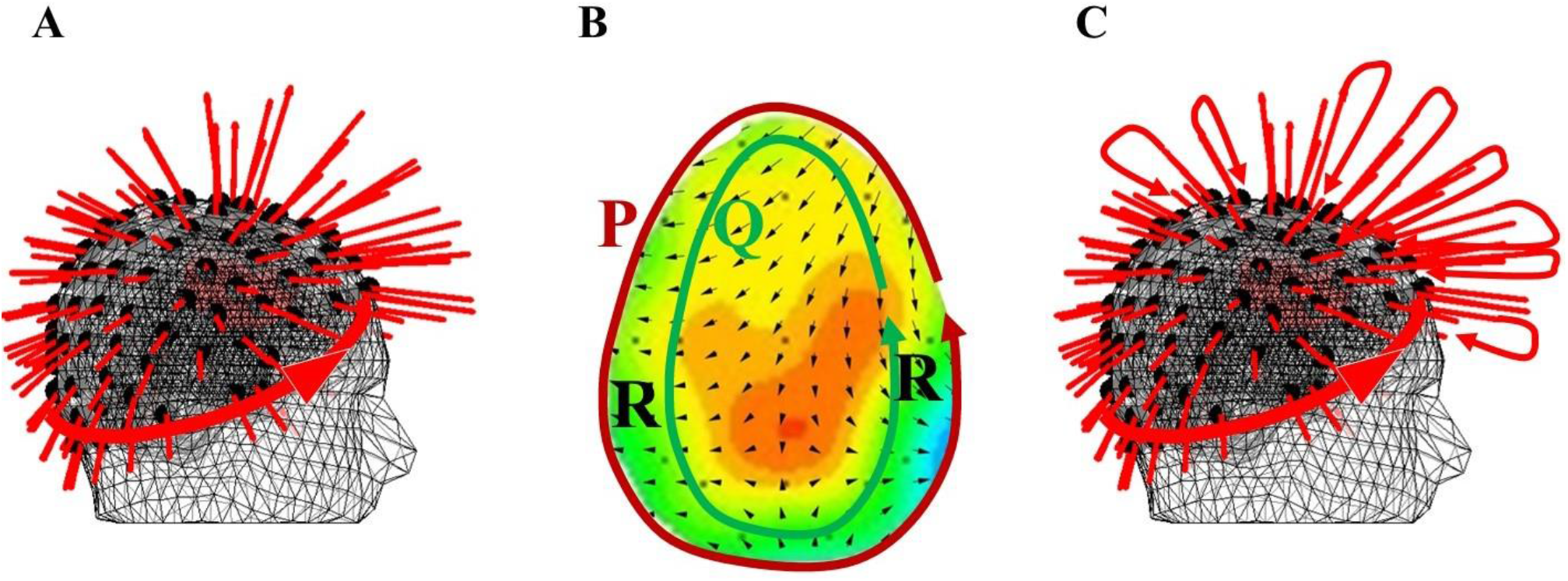
Orientations of the lead fields in the XYZ directions for headmodel dipoli for a superficial source. The single macroscopic value equalling the sum of all the microscopic vectorial activities (arrows) is outlined by the thick counterclockwise circle. Modified from: https://www.fieldtriptoolbox.org/workshop/oslo2019/forward_modeling/ **Figure 3B**. A pair of differentiable functions P and Q on an EEG oval. In the basic scenario described by the Green’s Theorem, P and Q are simple, closed curves that form the boundary R and the line integral C on Pdx + Qdy is a measurement of the area occupied by the region R. **Figure 3C**. The red arrows of Figure 3A can be described in terms of cycles and overlapping cycles standing for the vortexes formed by nested cycles.

The study of Green’s Theorem in terms of a piecewise smooth boundary formed by a pair differentiable functions P and Q is a source of a variety of applications (Zenisek, A., 1998; Tewari, R., Vinodchandran, N.V.). It has been proved that, whenever the following conditions for a pair of simple, closed curves (named after Jordan) are satisfied, then Green’s theorem holds (Craven, 1964).

When the simple (no loops), closed curves P and Q are close enough (**Figure 3B**), then the line integral 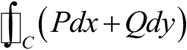 is approximately the same as the area of the region *R* within a piecewise smooth boundary traced by P and Q, i.e., whenever we have:

C = simple (not loops) Jordan Curve.

R = bounded region enclosed by the curve C (see **Figure 3C** for an example).

P(x,y), Q(x,y) = functions differentiable at all points of R and continuous on C + R.

Therefore,

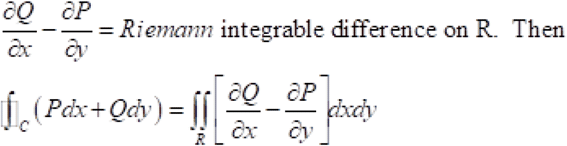

The conditions set forth by Craven (1964) for the Green’s theorem hold true with the pair of curves P, Q shown **in Figures 3B-3C**. Indeed, the cycles (simple, closed curves) traced by the differentiable functions P and Q are represented in the displays of brain waves. Obviously, P and Q together provide a boundary region R of the space occupied by the brain, viewed here in a planar fashion. In the case where 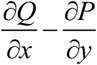 is an integrable difference, we know that Green’s theorem holds. From a practical standpoint, this means that Green’s theorem gives an approach to measuring the brainwave region bounded by P and Q in terms of the line integral 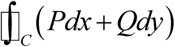, knowing that what we have found is the area of the region bounded by P and Q. In **Figure 3C**, this observation is applied to multiple bounded regions in pairs of overlapping cycles of brain activity vortexes. We now have a means of comparing the areas of bounded regions within different vortexes formed by nested cycles resulting from brain activity. These nested brain activity cycles are consistent with what we know about vortexes found in the nested, non-concentric cycles in electromagnetic wave forms (Baldomir, and Hammond, 1996).

To sum up, what do we achieve when using the Green’s theorem to evaluate nervous flows? A short answer to this question is: we arrive at a straightforward means of measuring the area of the region R bounded by a pair of nested curves commonly found in cortical activity vortexes. The collection of the vector fields detected from the scalp can be described by a single parameter, i.e., the counterclockwise electric flow occurring on the border of the EEG oval images. That is, we achieve glean information inside the shape traced by nested curves arising from brain activity.

## Data Availability

All data produced in the present study are available upon reasonable request to the authors

## ACKNOWLEDGMENT

To the memory of Norbert Jausovec. He and his precious EEG data go on living in our hearts and manuscripts.

